# The Role of Host Genetic Factors in Coronavirus Susceptibility: Review of Animal and Systematic Review of Human Literature

**DOI:** 10.1101/2020.05.30.20117788

**Authors:** Marissa LoPresti, David B. Beck, Priya Duggal, Derek A. T. Cummings, Benjamin D. Solomon

**Affiliations:** University of Florida College of Veterinary Medicine, Gainesville, Florida, United States of America; Inflammatory Disease Section, National Human Genome Research Institute, National Institutes of Health, Bethesda, Maryland, United States of America; Department of Epidemiology, Johns Hopkins Bloomberg School of Public Health, Baltimore, Maryland, United States of America; Department of Biology, University of Florida, Gainesville, Florida, United States of America; Emerging Pathogens Institute, University of Florida, Gainesville, Florida, United States of America; Office of the Clinical Director, National Human Genome Research Institute, National Institutes of Health, Bethesda, Maryland, United States America

**Keywords:** Coronavirus, COVID-19, Host genetic factors, SARS-CoV-2

## Abstract

**Background:** The recent SARS-CoV-2 pandemic raises many scientific and clinical questions. One set of questions involves host genetic factors that may affect disease susceptibility and pathogenesis. New work is emerging related to SARS-CoV-2; previous work on other coronaviruses in humans or other host species may be relevant.

**Objectives:** To review existing literature on host genetic factors and their association with infection and disease with coronaviruses in humans and in other host species.

**Methods:** We conducted a systematic review of literature on host genetic factors in humans associated with coronavirus outcomes. We also reviewed studies of host genetic factors associated with coronavirus outcomes in non-human species. We categorized articles, summarized themes related to animal studies, and extracted data from human studies for analyses.

**Results:** We identified 1,187 articles of potential relevance. Forty-five studies examined human host genetic factors related to coronavirus, of which 35 involved analysis of specific genes or loci; aside from one meta-analysis on respiratory infections, all were candidate-driven studies, typically investigating small numbers of research subjects and loci. Multiple significant loci were identified, including 16 related to susceptibility to coronavirus (of which 7 identified protective alleles), and 16 related to outcomes or clinical variables (of which 3 identified protective alleles). The types of cases and controls used varied considerably; four studies used traditional replication/validation cohorts. Of the other studies, 28 involved both human and non-human host genetic factors related to coronavirus, and 174 involved study of non-human (animal) host genetic factors related to coronavirus.

**Key findings:** We have outlined key genes and loci from animal and human host genetic studies that may bear investigation in the nascent host genetic factor studies of COVID-19. Previous human studies have been limited by relatively low numbers of eligible participants and limited availability of advanced genomic methods. These limitations may be less important to studies of SARS-CoV-2.

## Introduction

The ongoing severe acute respiratory syndrome coronavirus 2 (SARS-CoV-2) pandemic raises many scientific and clinical questions. One unknown is the extent to which individuals vary in susceptibility to infection and disease (COVID-19). Various hypotheses have been suggested to explain observed differences, including sex, age, comorbidities, and genetic factors.^1^ As with many complex diseases, the explanations likely involve a combination of genetic and non-genetic factors. In this context, genetic factors involve an interplay between virus and host genetics.^2^

Large, international studies and collaborations have formed to investigate host genetic factors related to COVID-19, including disease severity and susceptibility. These investigations include analyses of existing public and private datasets, as well as the establishment of new cohorts (e.g., https://blog.23andme.com/23andme-research/genetics-and-covid-19-severity/).^3^

While SARS-CoV-2 has seized recent attention, there are many other coronaviruses and a significant related body of literature exists about host genetic factors and their association with infection and outcomes in both humans and non-human host species. The *Coronavirinae* subfamily of the *Coronaviridae* family consists of four genera. The alphacoronaviruses include two major human coronaviruses, HCoV-229E (of which multiple HCoV-229E-like strains have been identified) and HCoV- NL63. Alphacoronaviruses that affect other species include mouse hepatitis virus (MHV), feline coronavirus (FCoV), which includes feline infectious peritonitis virus (FIPV) and feline enteric coronavirus (FECV), canine coronavirus (CCoV), and transmissible gastroenteritis coronavirus (TGEV) and porcine transmissible gastroenteritis coronavirus (TGEV) in pigs. The betacoronaviruses consist of four lineages: lineage A (HCoV-OC43 and HCoV-HKU1), lineage B (SARS-CoV-1 and SARS-CoV-2), lineage C (Middle East Respiratory Syndrome (MERS) and many bat coronaviruses), and lineage D (coronaviruses only identified in bats to date). HCoV-OC43, HCoV-229E, HCoV-HKU1, and HCoV-NL63 can result in a variety of presentations, including “common cold” and severe but rarely fatal disease; they are also frequently detected as co-infections with other viruses.^4^ There are other rare coronaviruses observed in humans as well as in other species.^5,6^ Relative to other coronaviruses, SARS-CoV-2 has unique biological properties and related clinical impact, but data regarding other coronaviruses may be relevant.

In various species, much work has focused on the genes encoding the relevant coronavirus receptor, including effects of viral and host genetic changes. Among other cell surface determinants,^7^ these receptor genes include *ACE2* for HCoV-NL63,^8^ SARS-CoV-1,^9,10^ and SARS-CoV-2,^11^ *ANPEP* for HCoV- 229,^12,13^ FIPV,^14^ CCoV,^15^ and TGEV,^16^ *DPP4* for MERS,^17–19^ and *Ceacam1* for MHV.^20^ Host genetic studies have - to varying degrees and in different ways - analyzed these genes, as well as other genes identified through targeted and agnostic methods. Studies to date have been disparate in terms of the virus and species studied, as well as the aims of the particular study. This has resulted in a rich body of literature that is difficult to efficiently leverage for SARS-CoV-2-related work.

We aimed to perform a review of the literature to outline previous studies of host genetic factors related to coronaviruses, with the objective of performing a systematic review to encapsulate genes and loci interrogated through these efforts. We do not attempt to fully describe the findings nor recapitulate what is known about the underlying host biology related to coronavirus infection and disease. As the majority of studies are candidate-driven, we did not attempt to conduct a metaanalysis. However, one goal is that the data presented here can help populate lists of genes that - along with data from related work^21–23^ - may bear scrutiny in the developing and important large-scale host genetic studies related to SARS-CoV-2.^24,25^ We present an overview of themes and interrogated genes/loci from animal studies, and perform a systematic review on human studies.

## Methods

We conducted an initial search of the PubMed database (last queried May 4, 2020) using each of the following phrases: “host genetics”; “genetic resistance”; “genetic susceptibility”; “genetic factors”; “genetics”; “GWAS” along with each of the following terms: “coronavirus”; “SARS”; “MERS”; “COVID- 19”; “COVID19”. We also identified additional articles by searching for specific coronaviruses or coronavirus-associated conditions (e.g., “canine coronavirus”; “middle east respiratory syndrome”) along with the term “genetics”. Articles were included in the search regardless of publication date. Articles included electronic, ahead-of-print publications available in the PubMed database. We also identified and categorized relevant articles from the references of initially selected articles. We did not include articles only available on non-peer reviewed preprint servers, though recognize that a substantial number of these manuscripts will be on PubMed soon.

Each abstract was reviewed by a single reviewer. Full articles were reviewed when insufficient data were available in the abstract, or when no abstract was available. Publications were classified into the following categories: 1) Study of human host genetic factors related to coronavirus; 2) Study of nonhuman (animal) host genetic factors related to coronavirus; 3) Study of non-genetic (including non-DNA-based analyses - see further explanation below) host factors related to coronavirus, including involving immunopathogenesis; 4) Study of other pathogens (not coronavirus); 5) Other studies of coronavirus. Articles containing information in both categories 1 and 2 were identified as such; articles were otherwise categorized according to the lowest numerical category (e.g., an article involving both human host genetic factors to coronavirus as well as immunopathogenesis would be categorized into group 1. Articles that did not involve investigations of specific DNA-based genetic changes (e.g., transcriptomic or proteomic studies) were categorized into group 3, as were studies that only included analyses of sex without other genetic analyses. Other publications, including: 6) Untranslated studies in another language (not English); 7) Not relevant (unrelated to coronavirus or other pathogens); 8) No data available; were removed from further analysis after categorization into these latter four categories. Data from category 1 publications were manually extracted for relevant information pertaining to: coronavirus studied; general methods and questions analyzed; gene(s), variant(s), or loci analyzed; size of cohorts studied; geographic or ancestral composition of cohorts; statistical results, including (where available) odds ratios, confidence intervals, and p-values.

## Results

Our search identified 1,187 articles of potential relevance (Figure 1, Supplementary Table 1). Of these, 45 involved study of human host genetic factors related to coronavirus (Table 1); 35 of the 45 human studies involved analysis of specific genes or loci (only one was a non-candidate study), while 10 involved biological, computational, or case report studies of human host genetic factors. Twenty-eight involved both human and non-human host genetic factors related to coronavirus (these largely investigated inter-species differences in disease susceptibility and pathogenesis, such as related to differences in *ACE2*); 174 involved study of non-human (animal) host genetic factors related to coronavirus; 584 involved study of non-genetic host factors related to coronavirus, including involving immunopathogenesis; 16 involved study of other pathogens (not coronavirus); 321 involved other studies of coronavirus. 18 studies were assigned to the other categories and removed.

**Figure 1.**
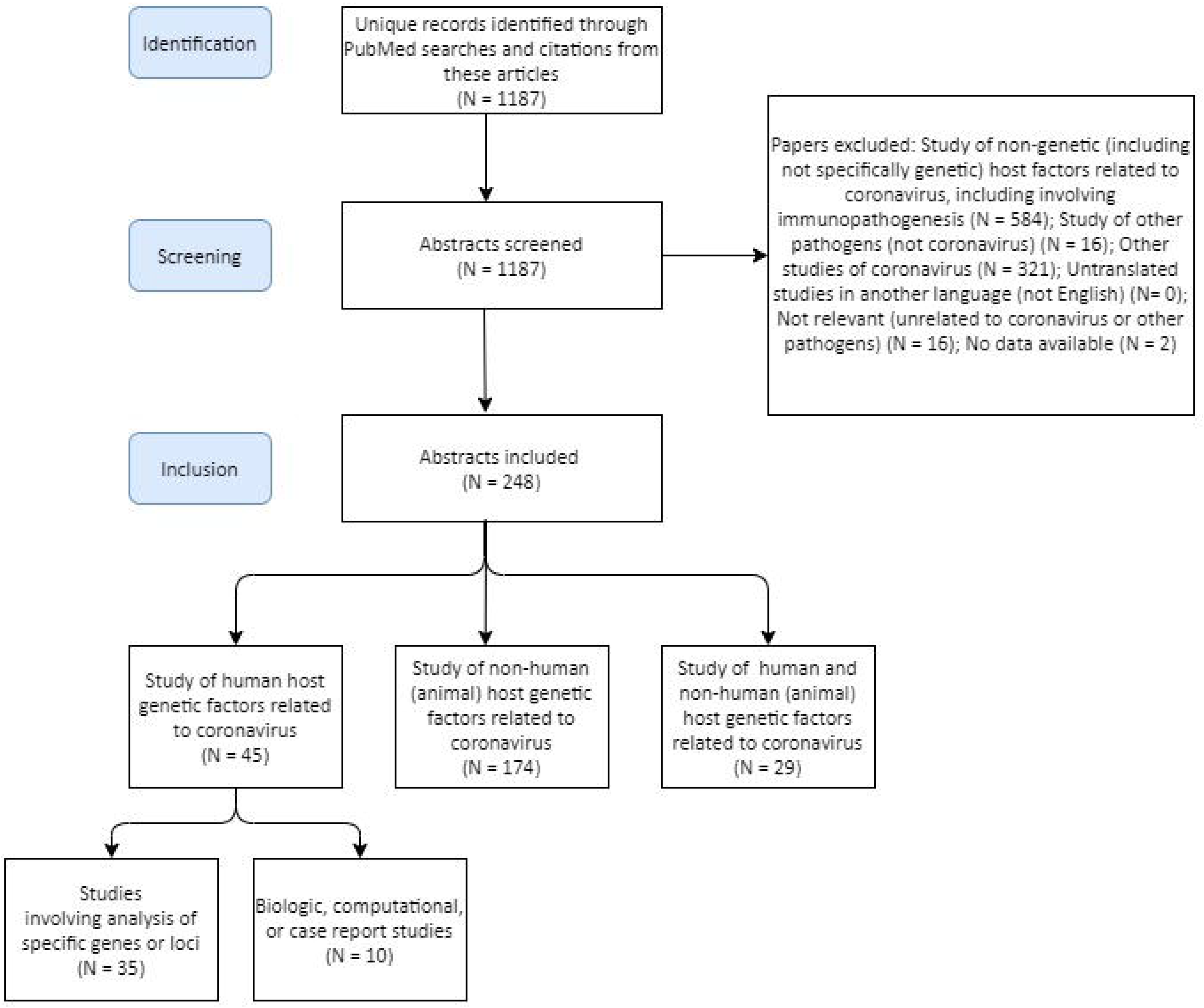
Description of articles identified through PubMed searches described in Methods.

**Table 1.** Summary of human studies (related to specific genes or loci) on host genetic factors related to coronaviruses. More details are available in Supplementary Table 2.

| Human | Method(s) or approach(es) | Key findings | PMID |
| --- | --- | --- | --- |

| coronavirus<br>studied (other<br>coronaviruses<br>or pathogens) |  |  |  |
| --- | --- | --- | --- |
| SARS-CoV-1 | Analysis of association of *HLA* gene polymorphisms with susceptibility to SARS-CoV-1 infection or clinical parameters | Association of HLA-B* 4601 with severity of SARS-CoV-1 infection | 12969506<sup>199</sup> |
| SARS-CoV-1 | Analysis of association of *HLA* gene polymorphisms with susceptibility to SARS-CoV-1 infection | HLA-B*0703, HLA-DRB1*0301 and co-inheritance of HLA-B*0703 and HLA-B60) were associated with susceptibility to SARS-CoV-1 infection | 15243926<sup>200</sup> |
| SARS-CoV-1 | Analysis of association of *ACE2* polymorphisms with SARS-CoV-1 clinical parameters | No association of *ACE2* polymorphisms with SARS-CoV-1 outcomes | 15331509<sup>201</sup> |
| SARS-CoV-1 | Analysis of association of *ACE* insertion/deletion (I/D) polymorphism with susceptibility to SARS-CoV-1 or clinical parameters | *ACE* D allele was associated with hypoxemia in SARS-CoV-1 infections | 15381116<sup>202</sup> |
| SARS-CoV-1 | Analysis of association of *OAS1*, *PKR*, *MX1* polymorphisms with susceptibility to SARS-CoV-1 or clinical parameters | *OAS1* rs3741981 and rs2660 were associated with SARS-CoV-1 susceptibility; *MX1* rs2071430 was associated in hypoxemia in SARS-CoV-1 infections | 15766558<sup>203</sup> |
| SARS-CoV-1 | Analysis of association of *ACE* insertion/deletion (I/D) polymorphism with susceptibility to SARS-CoV-1 or clinical parameters | No association was found with *ACE* insertion/deletion (I/D) polymorphism and susceptibility to SARS-CoV-1 or clinical parameters | 15819995<sup>175</sup> |
| SARS-CoV-1 | Analysis of association of *MBL* polymorphisms susceptibility to SARS-CoV-1 or clinical parameters and biological study of MBL | Serum MBL was lower in patients with SARS-CoV-1 infections than controls, and haplotypes associated with lower serum MBL were more frequent in patients with SARS-CoV-1 infections than in control subjects, but there was not association with mortality | 15838797<sup>204</sup> |
| SARS-CoV-1 | Analysis of association of *ACE2* polymorphisms and | No association was found with *ACE2* polymorphisms and | 15937940<sup>205</sup> |
|  | susceptibility to SARS-CoV-1 infection | susceptibility to SARS-CoV-1 infection |  |
| SARS-CoV-1 | Analysis of association of *MBL* polymorphisms and susceptibility to SARS-CoV-1 infection | *MBL* rs1800450 was associated with susceptibility to SARS-CoV-1 infection | 16170752<sup>206</sup> |
| SARS-CoV-1 | Analysis of association of *FCGR2A* and *MBL* polymorphisms and susceptibility to SARS-CoV-1 infection or clinical parameters | Homozygosity for *FCGR2A* rs1801274, as well as a linear trend of *FCGR2A* genotypes, was associated with severe SARS-CoV-1 infection | 16185324<sup>207</sup> |
| SARS-CoV-1 | Analysis of association of *CLEC4M* VNTR polymorphism with susceptibility to SARS-CoV-1 and biological studies of cells with these polymorphisms | Homozygosity for the *CLEC4M* VNTR polymorphism was associated with susceptibility to SARS-CoV-1, and homozygous cells had higher binding capacity for SARS-CoV-1, higher proteasome-dependent viral degradation, and lower capacity for trans infection. | 16369534<sup>208</sup> |
| SARS-CoV-1 | Analysis of association of *HLA* polymorphisms with SARS-CoV-1 susceptibility | HLA-Cw*0801 was associated with susceptibility to SARS-CoV-1 infection | 16455884<sup>209</sup> |
| SARS-CoV-1 | Analysis of association of polymorphisms in 65 genes with SARS-CoV-1 viral shedding | SARS-CoV-1 shedding was associated with alleles of *IL18*, *IL1A*, *RELB*, and *FLG2* | 16652313<sup>210</sup> |
| SARS-CoV-1 | Analysis of association of *OAS1* and *MX1* polymorphisms with susceptibility to SARS-CoV-1 | *OAS1* 3'-UTR rs2660 and *MX1* promoter rs2071430 were associated with susceptibility to SARS-CoV-1 | 16824203<sup>211</sup> |
| SARS-CoV-1 | Analysis of association of *CLEC4M* VNTR polymorphism with susceptibility to SARS-CoV-1 infection | No association was found with homozygosity for the *CLEC4M* VNTR polymorphism and susceptibility to SARS-CoV-1 | 17534354<sup>212</sup> |
| SARS-CoV-1 | Analysis of association of *CLEC4M* VNTR polymorphism with susceptibility to SARS- | No association was found with homozygosity for the *CLEC4M* VNTR polymorphism and | 17534355<sup>213</sup> |
|  | CoV-1 infection | susceptibility to SARS-CoV-1 |  |
| SARS-CoV-1 | Analysis of association of *CCL5*, *CXCL9*, and *CXCL10* polymorphisms with susceptibility to SARS-CoV-1 infection or clinical parameters | *CCL5* rs2107538 was associated with susceptibility to SARS-CoV-1 in one cohort and severe outcomes of SARS-CoV-1 infection in another cohort | 17540042<sup>214</sup> |
| SARS-CoV-1 | Analysis of association of *FCER2* and *ICAM3* polymorphisms with susceptibility to SARS-CoV-1 or clinical parameters | Homozygosity for *ICAM* rs2304237 was associated with higher LDH levels and lower total WBC counts | 17570115<sup>215</sup> |
| SARS-CoV-1 | Analysis of association of *CD14*, *TLR2*, and *TLR4* polymorphisms with susceptibility to SARS-CoV-1 or clinical parameters | *CD14* rs2569190 was associated with severe SARS-CoV-1 infection (this data was also combined with previous data, suggesting that this and *FCGR2A*-RR131 are risk genotypes for severe SARS-CoV-1 infection) | 17913858<sup>216</sup> |
| SARS-CoV-1 | Analysis of association of *TNF* polymorphisms with | *TNF* polymorphisms were associated with susceptibility to | 18312678<sup>217</sup> |
|  | interstitial lung fibrosis and femoral head osteonecrosis in discharged SARS-CoV-1 patients | SARS-CoV-1 and with femoral head necrosis in discharged SARS-CoV-1 patients |  |
| SARS-CoV-1 | Analysis of association of polymorphisms in *IL12RB1* with susceptibility to SARS-CoV-1 or clinical outcomes | IL12RB1(+1664) polymorphism was associated with susceptibility to SARS-CoV-1 infection | 18478121<sup>218</sup> |
| SARS-CoV-1 | Analysis of association of polymorphisms in 4 C-type lectin genes with susceptibility to SARS-CoV-1 infection | No association of polymorphisms in C-type lectin genes with SARS-CoV-1 susceptibility | 18697825<sup>219</sup> |
| SARS-CoV-1 | Analysis of association of polymorphisms in 9 inflammatory response genes with susceptibility to SARS-CoV-1 or clinical outcomes | No association of polymorphisms in inflammatory response genes with SARS-CoV-1 susceptibility or clinical outcomes | 18708672<sup>220</sup> |
| SARS-CoV-1 | Analysis of association of polymorphisms in *MASP2* with susceptibility to SARS-CoV-1 infection | No association of *MASP2* polymorphisms with SARS-CoV-1 susceptibility | 19405982<sup>221</sup> |
| SARS-CoV-1 | Analysis of association of *HLA* polymorphisms with SARS-CoV-1 susceptibility | HLA-DRB1*12 was more frequent in SARS-CoV-1 patients versus controls; HLA-DRB1*1202 showed the strongest association with SARS-CoV-1 infection in a dominant model | 19445991<sup>222</sup> |
| SARS-CoV-1 | Analysis of association of polymorphisms in 64 genes with susceptibility to SARS-CoV-1 infection | CXCL10(-938AA) is protective (but appears jointly with other variants); *FGL2*(+158T/*) is associated with higher susceptibility unless combined with CXCL10/(-938AA), when jointly is associated with lower susceptibility | 19590927<sup>223</sup> |
| SARS-CoV-1 | Analysis of association of *CD209* rs4804803 with SARS-CoV-1 outcomes | *CD209* polymorphism rs4804803 is associated with lower LDH levels (and therefore, worse prognosis) | 20359516<sup>224</sup> |
| SARS-CoV-1 | Biological study and analysis of *MX1* promoter polymorphisms with suppressed interferon beta induction and association of *MX1* promoter polymorphisms with susceptibility to SARS-CoV-1 infection | Differences were observed in binding affinity to nuclear proteins related to IFN-beta stimulation; *MX1* rs2071430 was associated with lower risk of SARS-CoV-1 infection | 20462354<sup>225</sup> |
| SARS-CoV-1 | Analysis of association of *HLA* gene polymorphisms with SARS-CoV-1 susceptibility | No significant associations (after correction) *HLA* gene polymorphisms with SARS-CoV-1 susceptibility were identified | 20864745<sup>226</sup> |
| SARS-CoV-1 | Biological study of in vitro functional effects of rs4804803 and analysis of association of *CD209* rs4804803 with SARS-CoV-1 outcomes | *CD209* polymorphism rs4804803 was associated with lower risk of high admission LDH levels, and may contribute to a reduced immune response/reduced lung injury during disease progression | 20864747<sup>227</sup> |
| SARS-CoV-1 | Analysis of association of *AHSG* and *CYP4F3A* polymorphisms with SARS-CoV-1 susceptibility | *AHSG* polymorphism rs2248690 was associated with SARS-CoV-1 susceptibility (as well as higher | 21904596<sup>228</sup> |
|  |  | AHSG serum concentration) |  |
| SARS-CoV-1 | Analysis of association of *HLA* polymorphisms with SARS-CoV-1 susceptibility | *HLA*-Cw*1502 conferred resistance against SARS infection is associated with resistance to SARS-CoV-1 infection | 21958371<sup>229</sup> |
| SARS-CoV-1 | Analysis of association of *HLA* polymorphisms with SARS-CoV-1 susceptibility and outcome | No association of *HLA* polymorphisms with SARS-CoV-1 susceptibility and outcome were identified | 24643938<sup>230</sup> |
| SARS-CoV-1 | Analysis of association of *CCL2* and *MBL* polymorphisms with susceptibility to SARS-CoV-1 infection | Variants in *MBL* (rs1800450) and *CCL2* (rs1024611) (*CCL2*) were cumulatively associated with SARS-CoV-1 susceptibility | 25818534<sup>231</sup> |
| SARS-CoV-1 (and other respiratory pathogens) | Meta-analysis of 386 studies on susceptibility to tuberculosis, influenza, respiratory syncytial virus, SARS-CoV-1, and pneumonia | In a pooled model, variants in *IL4* were positively associated with susceptibility after multiple testing correction | 26524966<sup>174</sup> |
| SARS-CoV-2 | Case report of death due to | Suggestion of genetic | 32277694<sup>232</sup> |
|  | COVID-19 in three previously healthy adult brothers | predisposition due to apparent familial clustering |  |
| SARS-CoV-2 | Case reports of two patients with X-linked agammaglobulinemia (and documented pathogenic variants in *BTK*) | Patients recovered, suggesting that B cell response might not be required to overcome the SARS-CoV-2 infection | 32319118<sup>233</sup> |
| SARS-CoV-2 | Analysis of association of *IFITM3* rs12252 with clinical outcomes of SARS-CoV-2 infection | Significant association of homozygosity *IFITM3* rs12252 with disease severity | 32348495<sup>234</sup> |

Abbreviations: CCoV: canine coronavirus; FCoV: feline coronavirus; human coronavirus 229E: HCoV-229E; human coronavirus NL63: HCoV NL63; human coronavirus OC43: HCoV OC43; LDH: lactate-dehydrogenase; MBL: Mannose-binding lectin; MERS-CoV: middle east respiratory syndrome coronavirus; SARS-CoV-1: severe acute respiratory syndrome coronavirus 1; SARS-CoV-2: severe acute respiratory syndrome coronavirus 2; SL-CoV: SARS-Cov-1-like coronaviruses; TGEV: porcine transmissible gastroenteritis coronavirus; WBC: white blood cell; WT: wild-type

We organized our analysis and findings into the schema presented below.

### Animal studies

Coronaviruses affect many species, from Beluga whales to spotted hyenas to turkeys, and sequelae of disease can range from apparently asymptomatic infections to severe or lethal effects on different organ systems, potentially manifesting as diarrheal, encephalitic, nephritic, respiratory, and other types of disease.^26,27^ There are numerous non-observational animal studies of coronaviruses, such as involving hamsters,^28–30^ guinea pigs,^31^ rats,^10,32–35^ and non-human primates.^36–38^ However, formal host genetic studies have been described for some but not all species. Many studies have involved examination of differences in species susceptibility and pathogenesis to human and non-human coronaviruses.^14,39–41^

Among the host genetic studies in animals, the objectives and methods used differ significantly depending on the species studied. For example, in chickens and other livestock, the types of published studies predictably differ from those conducted on experimental mice. That is, while MHV represents a problem for mouse colonies, the rationale of the livestock studies may focus more purely on economic repercussions versus attempts to use a model organism to understand immunopathogenesis of infectious disease.^42^ The degree to which results may be reported through the scientific literature (versus other routes) is also anticipated to differ depending on the species studied and the reason for the study. See Figure 2 for a summary of interrogated loci in animal studies.

**Figure 2.**
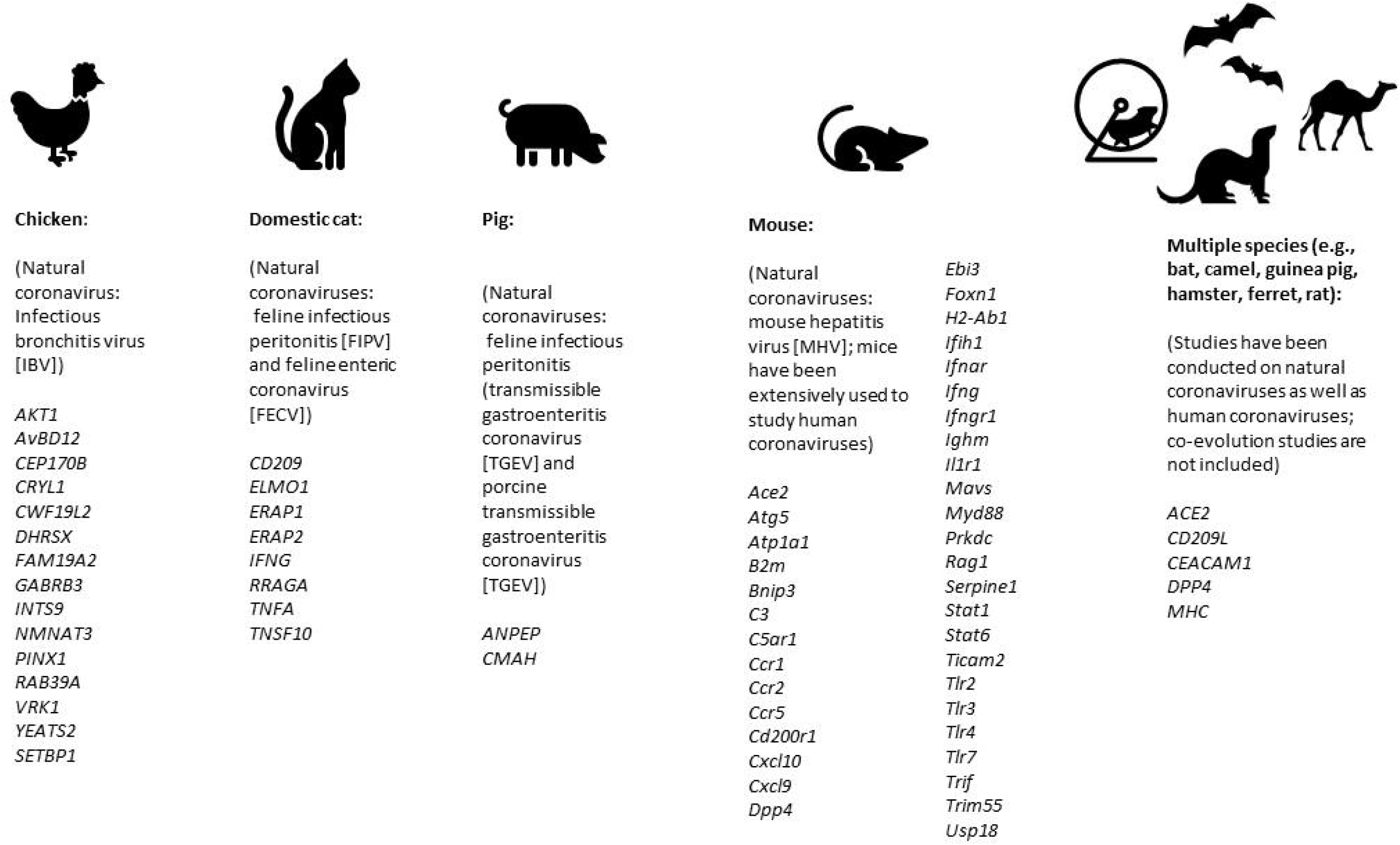
Genes investigated in animal studies related to coronavirus disease. Human genes are shown only for those studies in multiple species analyses; other human gene details are presented elsewhere.

We describe representative studies and key findings below, but the descriptions should not be considered as truly comprehensive; additionally, as noted above, many studies compared susceptibility across species, both through cell-based assays and experimental animals. Many investigations using other methods (e.g., transcriptomics or proteomics) have identified key molecules involved in coronavirus susceptibility and pathogenesis. Though beyond the scope of this article, these molecules should also be considered in future SARS-CoV-2 host genetic studies.

### Model animal strains, experimental animals, and domesticated animals

#### Chicken

In chickens, the infectious bronchitis virus (IBV) coronavirus can cause disease affecting different organ systems and tissues, such as IBV-associated nephritis. As with other species, inbred status and specific chicken lines impact host susceptibility, immune response, and outcomes, and virus/host genetic interactions have been described.^43–47^ Breeding experiments have suggested different inheritance patterns related to susceptibility and outcomes, and have implicated both MHC and non-MHC loci.^48,49^ Multiple GWAS investigating the immune response to IBV have identified significantly-associated polymorphisms in the breeds studied;^50,51^ the implicated or nearest genes include: *AKT1, AvBD12, CEP170B, CRYL1, CWF19L2, DHRSX, FAM19A2, GABRB3, INTS9, NMNAT3, PINX1, RAB39A, VRK1, YEATS2;* and *SETBP1*:^50,51^

#### Domestic cat

Felines can be infected by feline coronavirus (FCoV), which include feline infectious peritonitis (FIPV) and feline enteric coronavirus (FECV).^52^ As with other species, cats demonstrate a range of potential effects. In addition to association with traits such as age, sex, and reproductive status, purebred status and loss of heterozygosity has been shown to be associated with the effects of disease. Susceptibility and outcomes also appear to vary between different breeds.^52–60^ A small study of feline leukocyte antigen (FLA)-DRB alleles did not show a statistically significant association between the number of FLA-DRB alleles and FCoV infection outcome.^61^ Polymorphisms in *IFNG* (investigated as FIP can result in decreased interferon-gamma levels) were shown to correlate with plasma interferon-gamma levels and outcomes.^62^ Polymorphisms in *TNFA* and *CD209* were also shown to be associated with outcomes in one inbred line.^63^

In addition to candidate studies, several GWAS have been performed in cats. One small study on outcomes in experimentally-induced infections in random-bred cats identified one associated genomic region (which did not harbor any obvious candidate genes).^52^ Another small study on an inbred breed identified multiple candidate genes *(ELMO1, ERAP1, ERAP2, RRAGA, TNSF10)* but none was fully concordant with the FIP disease phenotype.^64^ Recent studies on SARS-CoV-1 and SARS-CoV-2 have investigated the susceptibility of cats as well as other animals;^65^ see further details below (under Ferrets).

#### Dromedary camel

Camels are an important reservoir of coronaviruses that can infect humans; this became especially relevant in the context of MERS.^66–68^ Many studies have analyzed factors that contribute to spread,^69^ though the searches employed in this analysis identified relatively few host genetic studies separate from analyses of DPP4 receptor characteristics and tropism, including comparisons between camels, humans, and other species.^70–74^

#### Ferret

Several studies have investigated the susceptibility of various species to coronaviruses. One objective relates to identifying useful animal models of disease, in which non-human species show similar infection and disease outcomes to humans upon exposure to coronaviruses.^65,75,76^ For example bat, camel, and humans can be infected by MERS, unlike mouse, ferret, hamster, and guinea pig. SARS-CoV- 2 replicates better in ferrets and cats than in dogs, pigs, chickens, and ducks. One explanation involves genetic characteristics of the host receptor for the relevant virus.^76,77^ Additionally, within an infected animal, the site of viral replication appears to vary according to the species and coronavirus, and is additionally potentially related to tissue-specific receptor expression.^78^ This line of reasoning may also be relevant to age-specific differences observed with SARS-CoV-2 and human infections.^79^

#### Hamster

As noted above, hamsters have been used as model organisms to study coronaviruses, including studies of host receptors. This includes studies using standard hamster cell lines as well as other approaches involving hamster models.^80–85^ For example, hamster models have been used to study species susceptibility to MHV (related to *Ceacam1*),^86^ how alterations of specific Dpp4 amino acids in hamster affect susceptibility to MERS,^71,87^ and the roles of ACE2 and CD209L in SARS-CoV-1 susceptibility.^82^

#### Mouse

MHV has represented a challenge for the health of mouse colonies, though relatively recent improvements in animal care practices have been beneficial.^88^ Differences in the susceptibility of different mouse strains to MHV has been noted for seven decades.^89–91^ Studies have examined a number of different MHV strains. These strains demonstrate different tissue tropism and have different effects on various mouse lines.^92^ One distinct example is the JHM strain of MHV, which causes encephalitis in susceptible animals.^93,94^ In the discussion below, though susceptibility and outcome findings will be summarized, it is important to note that studies generally focus on the interactions between certain MHV strains and mouse lines, and it is not always clear how well these findings extrapolate to other strains and lines.

Many studies have investigated biological explanations for differences in MHV susceptibility and pathogenesis.^95–97^ Studies examining different laboratory mouse strains have suggested that multiple loci are involved.^98–111^ Early studies suggested various models, including potential monogenic/Mendelian explanations as well as more complex explanations involving interacting loci.^92,112–115^

Among many studies aiming to understand the underlying pathophysiology, mouse studies originally focused on strains believed to be involved in host susceptibility and reaction to infection. Importantly, these studies have identified interactions of host genetic factors with other factors, such as the cellular environment,^116,117^ cell and tissue-specific effects related to viral as well as host genetics,^118–123^ and host age.^124–126^ Unsurprisingly, some aspects of the disease process appear to be independent of observed strain differences.^127^ These studies also showed that host genetic factors influence different parts of the disease process, from initial virus-receptor binding,^117^ to cellular viral spreading^128,129^ and multiple aspects of the immune response.^101,130–134^ These studies enabled the cloning of *Ceacam1*, the MHV receptor gene,^81^ as well as related work regarding how genetic changes affecting this receptor confers MHV resistance in SJL mouse lines via inhibition of viral integration into host cells.^103,106,135,136^

In addition to the above studies, MHV-based mouse studies have used transgenic models to directly test the role of implicated pathways (summarized in Table 2). Not surprisingly, the majority of work in mouse models have focused on pathways already implicated in viral infection susceptibility including adaptive immune responses including both humoral and cellular, specific cytokine and immune receptor pathways, viral receptors, complement pathway, apoptosis, autophagy, and tissue repair. These studies have prominently implicated Type I (β) and II (γ) interferon responses in host response and predominantly protection against MHV infection. However, not all pro-inflammatory pathways are protective. For example, complement activation promotes tissue damage caused by MHV infection, highlighting the complex interplay between the host and virus. In addition to targeted gene disruptions described above, a GWAS using a recombinant inbred mouse panel implicated *Trim55*, which is involved in vascular cuffing and inflammation in response to SARS-CoV-1.^137^

**Table 2.**
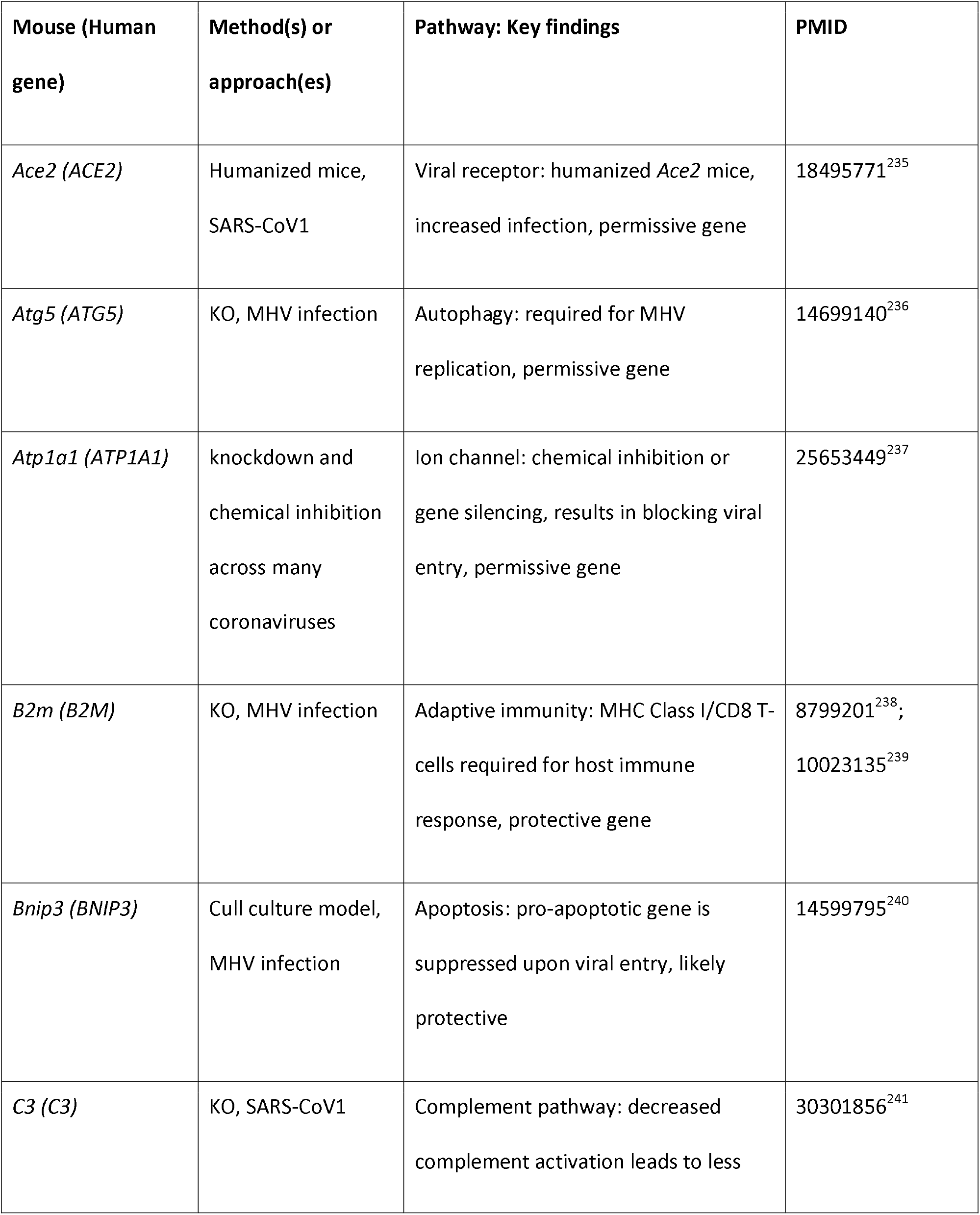
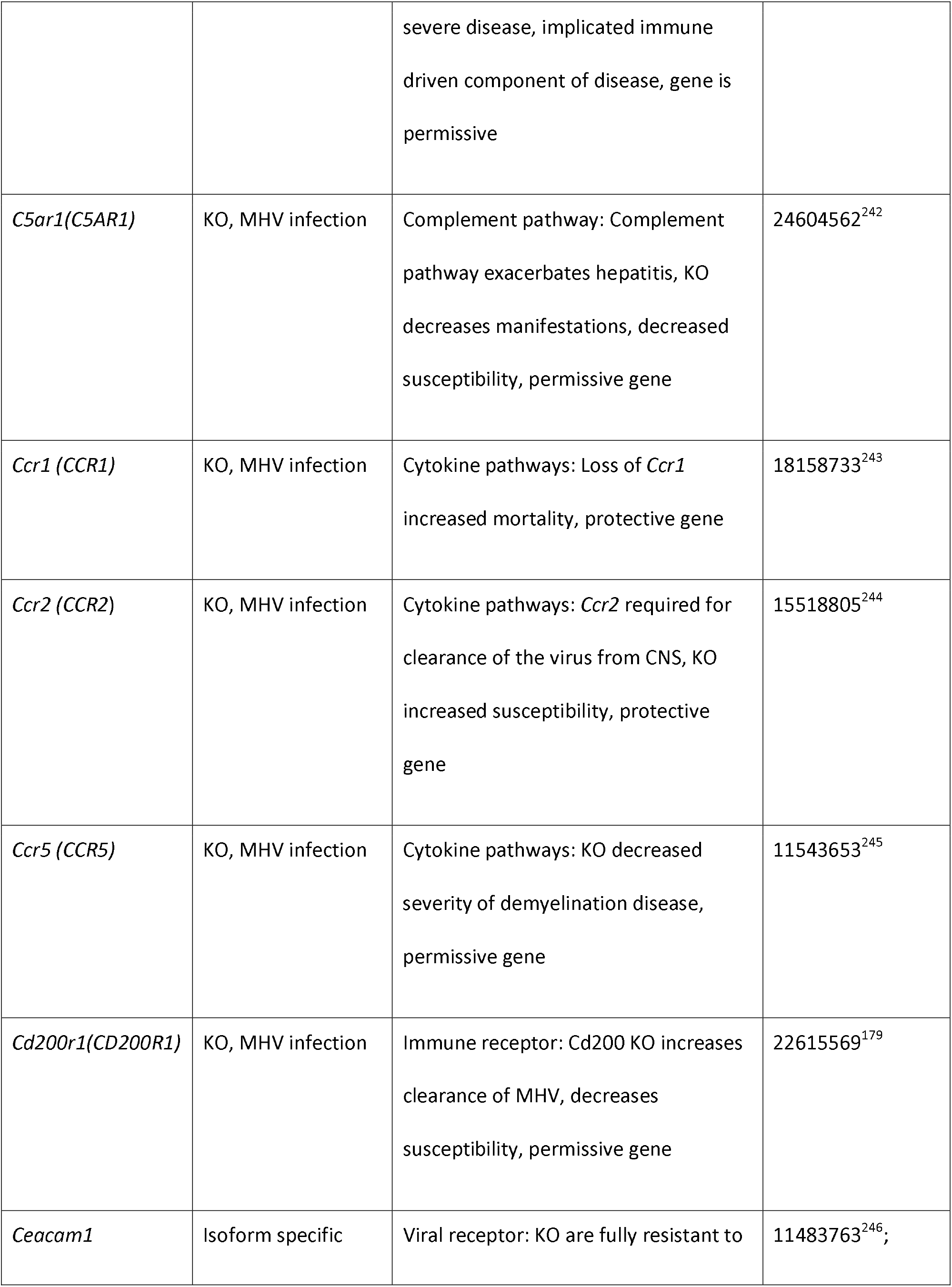
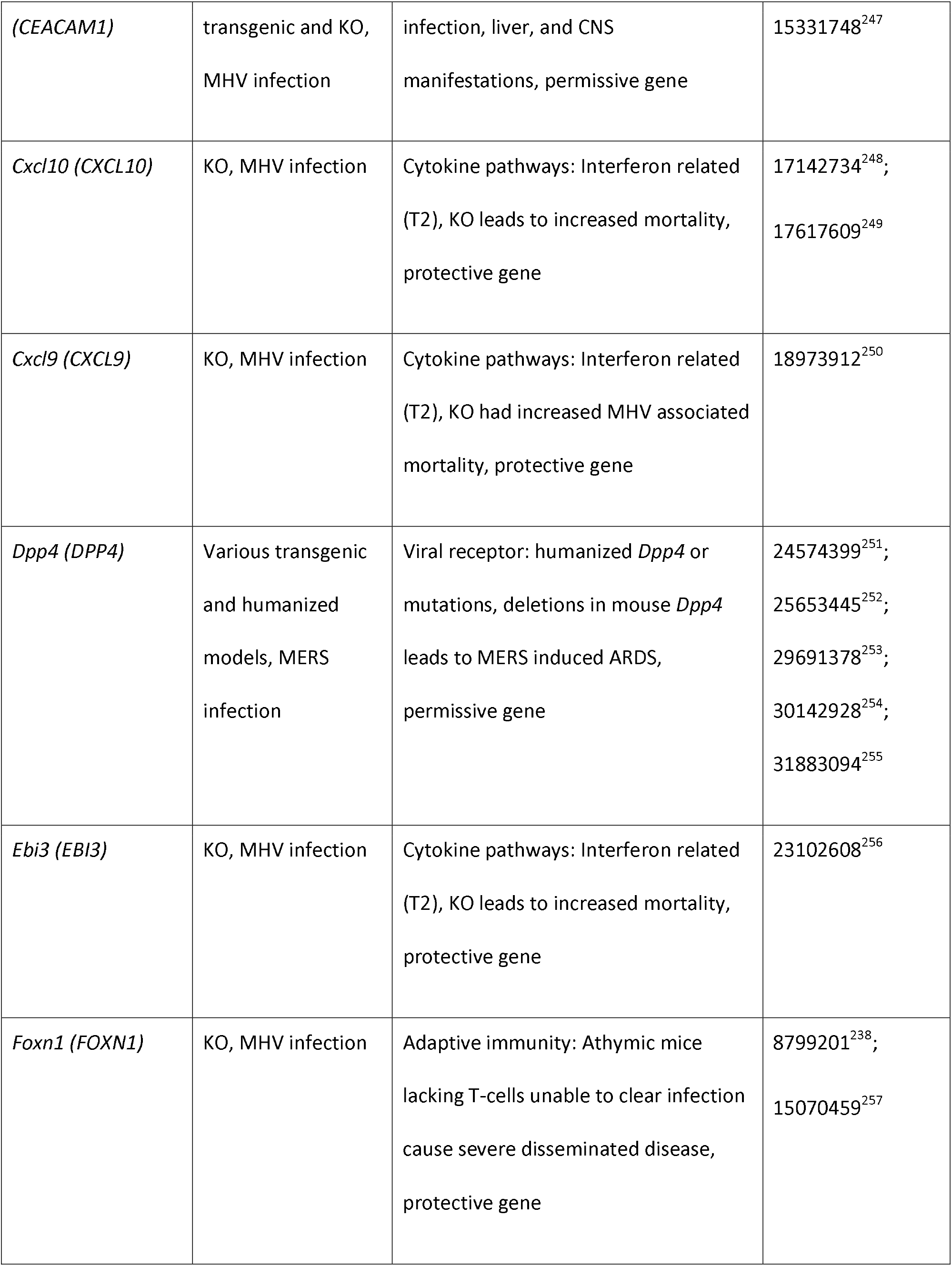
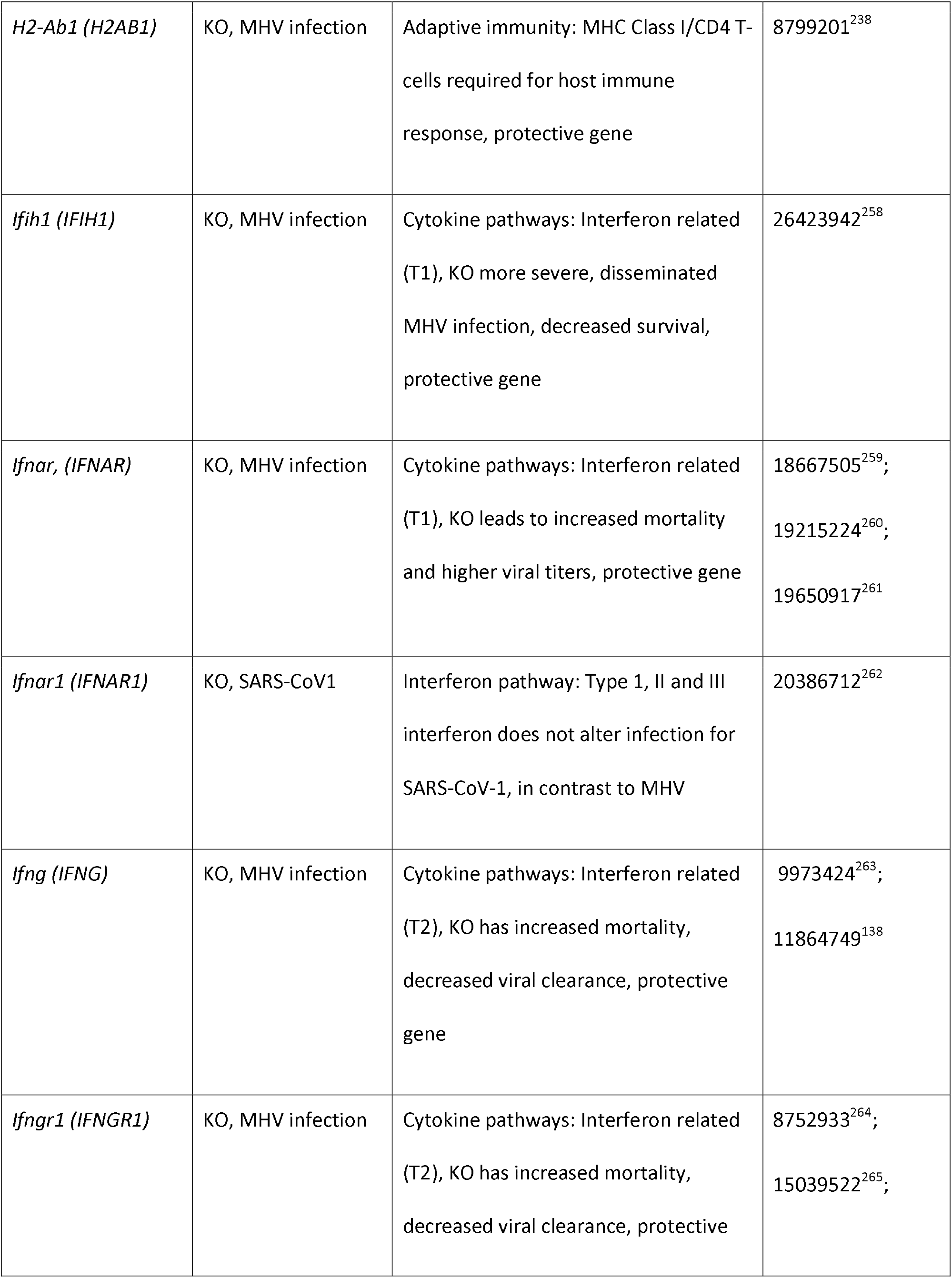
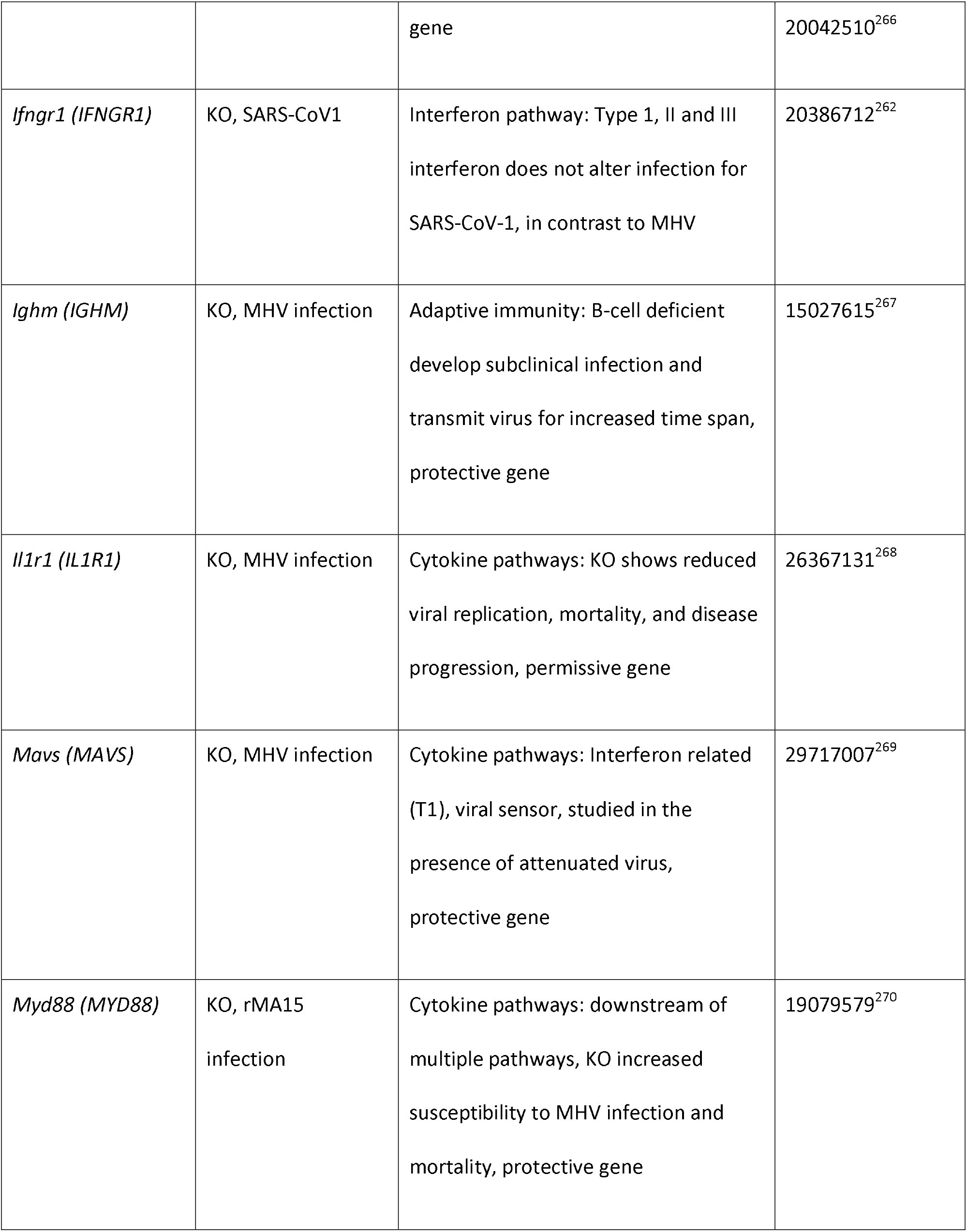
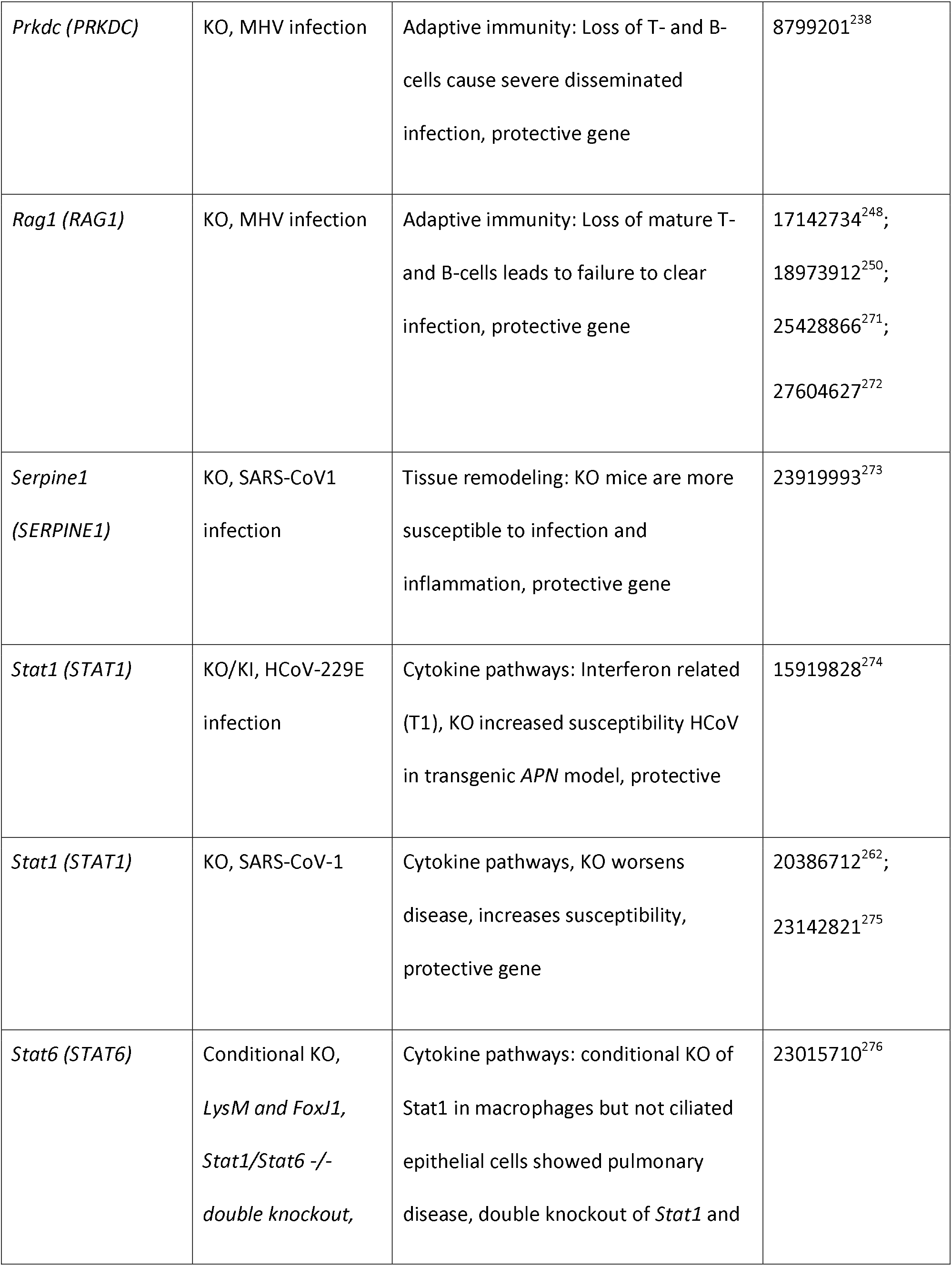
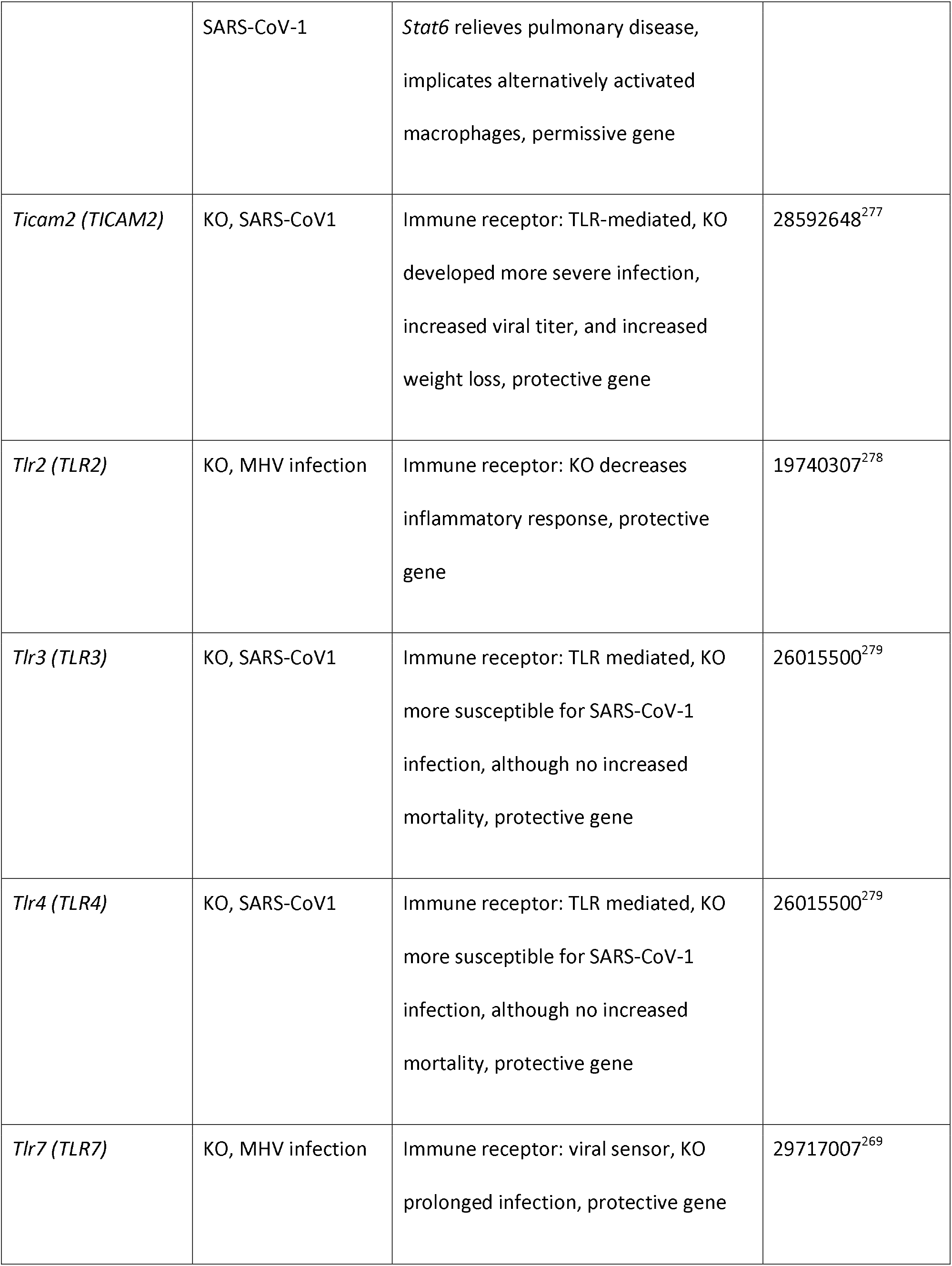
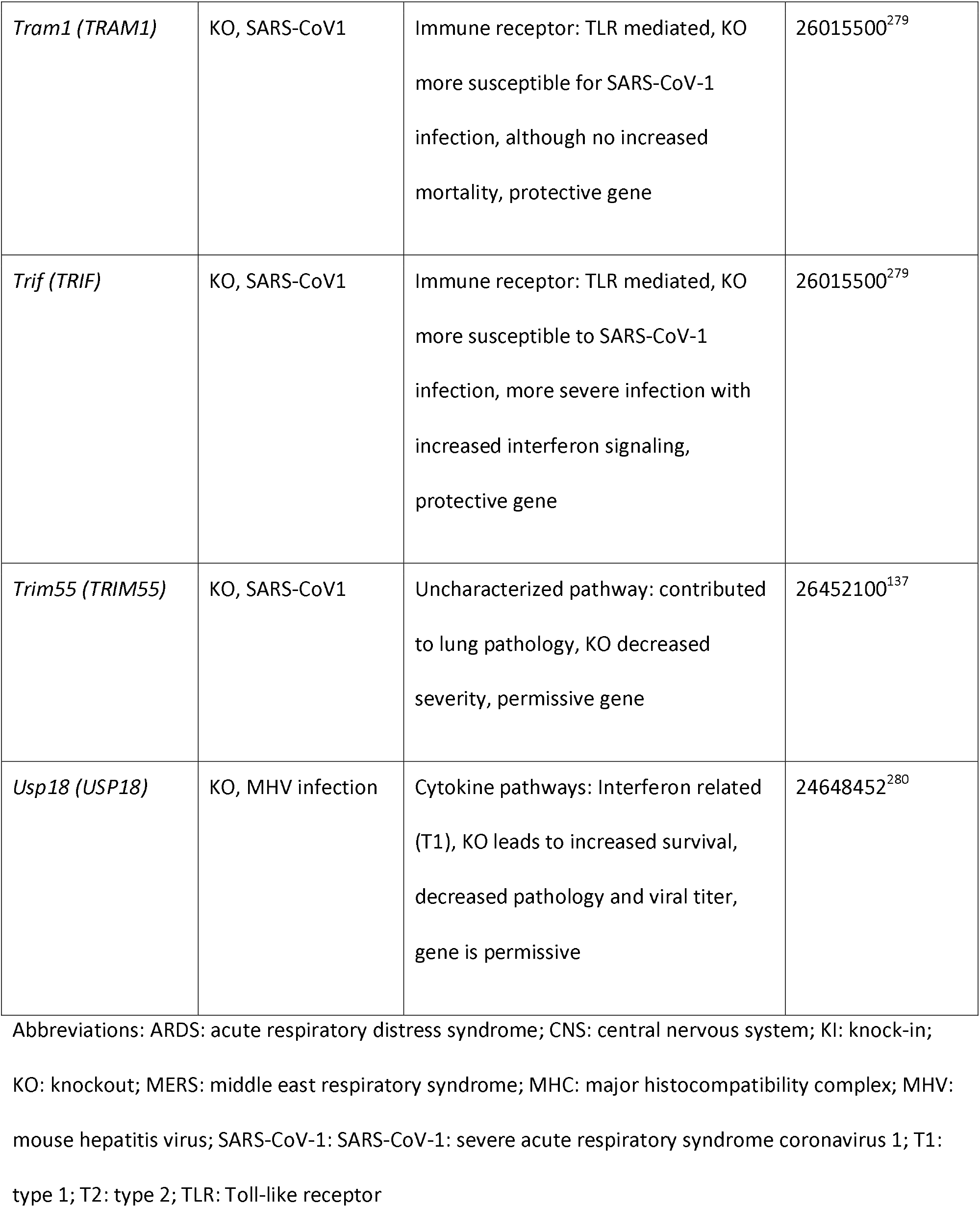
Summary of relevant mouse studies related to coronavirus. Note that the different studies have disparate objectives, many of which more directly involve aspects of immunopathogenesis versus standard host genetic questions regarding why specific genetic variants may affect disease susceptibility and outcomes.

Additional transgenic studies have investigated multiple biologic effects as well as returning to questions regarding susceptibility of different strains.^138^ Other mouse models (including knockouts, specific knockin mutations, humanized mice, and other models involving genetic manipulation) have been used to study human pathogens such as SARS-CoV and MERS; revealing similar properties for viral receptors, *Dpp4* for MERS, *Ace2* for SARS-CoV, cytokine and immune receptor pathways, and complement pathway as with mouse models of MHV. Intriguingly, there are differences between the importance of interferon pathways in host response to SARS-CoV1, where these pathways are dispensable as compared to MHV, where they are protective. Together, these different pathogen models have shown overlapping and unique pathways of host response between coronaviruses and highlight the potential relevance for SARS-CoV-2. See also the *Additional papers on humans and other species* section regarding further examples of studies involving mice and humans, as well as other species.

#### Pigs

Pigs can be infected by transmissible gastroenteritis virus (TGEV) and porcine epidemic diarrhea virus (PEDV), as well as the more recently-identified porcine deltacoronavirus (PDCoV). Like coronavirus disease in chickens, these diseases can have economic effects on the food industry,^139^ and analyses aim to address ways to ameliorate disease, such as the development of vaccines. Importantly, variants (both natural and experimentally-induced) may have different effects on different coronaviruses. For example, aminopeptidase N, encoded by *ANPEP* (also called *APN*) was reported as a functional receptor for TGEV and PEDV (as well as HCoV-229E), but multiple models, including CRISPR/Cas9-generated knock-outs, show differences in cellular susceptibility to TGEV and PEDV.^^139^,^140^^ In another study, infection by PEDV and TGEV correlated positively with ANPEP expression, but PEDV and TGEV could infect ANPEP-positive and negative enterocytes, with differences observed between viral strains.

Overall, the results suggest the presence of an additional receptor.^141^ Building on this type of work, site-specific editing *of ANPEP* has been suggested as a potential means to breed resistant animals.^142^ Studies focusing on PEDV have shown that knock-out of *CMAH* (hypothesized to affect cellular binding) does not result in immunity, but may improve outcomes.^143^

#### Rats

Rats can be affected by rat coronaviruses, and can be hosts to a number of different coronaviruses that affect other species.^144,145^ Rats have been used as model systems to investigate MHV, including through cellular-based assays.^146,147^ Several studies have examined rat susceptibility to various coronaviruses. As with many other studies, these have implicated key interactions between viral and host genetics that affect species and tissue tropism [17151094].^33^ In addition to computational approaches examining receptor characteristics, such as involving ACE2 in the context of SARS-CoV-1^10^ experimental studies suggest that rats are not susceptible to MERS based on *Dpp4* characteristics.^148^

### Non-domesticated animals

As described, many species can be infected by coronaviruses. These species include wild as well as domesticated animals. The below section provides select examples of genetic studies on wild animals. Others studies been conducted on coronaviruses (as well as other pathogens),^149^ especially related to host ranges or reservoirs and involving host/pathogen co-evolution.^150,151^ Related to host genetic studies that are particularly relevant to the current SARS-CoV-2 pandemic (e.g., pangolin), our searches did not identify relevant articles.

#### Cheetah

Among wild animals, severe population bottlenecks (resulting in reduced genetic diversity) in cheetahs has been used to explain their increased susceptibility to infection by FIPV as well as other infectious diseases. Several such bottlenecks appear to have occurred in cheetah, due to a combination of factors.^152–154^ Among possible explanations for this susceptibility, genetic uniformity of the major histocompatibility complex (MHC) has been suspected to be involved.^155^

#### Civet

Studies have focused on palm civets (as well as other species) related to zoonotic implications as this species has been implicated as the reservoir associated with introduction of SARS-CoV-1 into humans. Specifically, questions about host receptor characteristics (*ACE2*) have been described in the context of SARS-CoV-1.^156–158^ As with other coronaviruses and species, the interactions of viral and host genetics have been shown to be important.^159,160^

#### Bat

As a natural reservoir for many coronaviruses, bats have been studied more extensively than other species outside of laboratory-based animals and livestock. Studies have included co-evolutionary studies between coronaviruses and the genomes of bat hosts (e.g., by correlating phylogenetic analyses of bat coronaviruses with *CYTB* in multiple bat species)^161^ as well as genetic/biologic studies related to host genetic factors. These have involved well-studied genes such as *ACE2* with SARS-CoV-1^162,163^ and *DPP4* with MERS.^164,165^ In addition to allowing analyses of host susceptibility, these and similar studies help provide estimates for the time-frame of coronavirus circulation in species and populations, and explore cross-species transmission.^150^

## Human

Details of the human studies are presented in Table 1, Figure 3, and Supplementary Table 2. Forty-five studies were initially identified by the methods described. Of these, 35 involved association or other studies related to human host genetic factors (see summary in the next paragraph). Ten others involved biological, computational, or other non-genetic association studies. Many other studies were identified that used a combination of human and animal models, but were categorized separately; additionally, many studies that might be considered genetic studies - if the definition were applied less stringently - were grouped in category 3. For example, studies have examined how specific genes are involved in aspects of viral disease but did not strictly study how DNA-based host genetic variants affect this process. In summary, these ten included mapping of a susceptibility locus to HCoV-229E to chromosome 15.^166^ Multiple studies examined the biological effects of mutant genes. Studying the effects of mutant *ACE2* on SARS-CoV-1 entry provided evidence that the cytoplasmic tail of ACE2 is not required for SARS-CoV-1 penetration.^167^ Studies of mutant *TRIM56* on antiviral activity against HCoV-OC43 and other viruses showed that anti-HCoV-OC43 activity relies solely upon TRIM56 E3 ligase activity; this appears different from the mechanisms related to other viral pathogens.^168^ Knockout culture cells and nonsynonymous variant *PPIA* models result in limitation of HCoV-229E replication.^169^ (Please note that we did not separately or exhaustively investigate human genetic experiments involving cell culture systems.) Specific variants in *IFITM* genes (*IFITM1* and *IFITM3* were studied) modulate the entry of multiple human coronaviruses (HCoV-229E; HCoV-NL63; HCoV OC43; MERS-CoV; and SARS-CoV- 1 were studied).^170^ Computational models suggest that, while most *ACE2* variants have similar binding affinity for SARS-CoV-2 spike protein, certain variants (rs73635825 and rs143936283) demonstrate different intermolecular interactions with the spike protein.^171^ An in silico analysis of viral peptide-MHC class I binding affinity related to HLA genotypes for SARS-CoV-2 peptides, as well as potential cross-protective immunity related to four common human coronaviruses, provides evidence that HLA-B*46:01 may be associated COVID-19 vulnerability, while HLA-B*15:03 may enable cross-protective T-cell based immunity.^172^ A recent study on viral cell entry showed that SARS-CoV-2 uses ACE2 for cell entry and TMPRSS2 for S protein priming; potential interventions based on these results include TMPRSS2 inhibition and convalescent sera.^173^ In addition to these examples, there are undoubtedly other biological, computational, and other studies examining how changes in and affecting key proteins may modulate disease.

**Figure 3.**
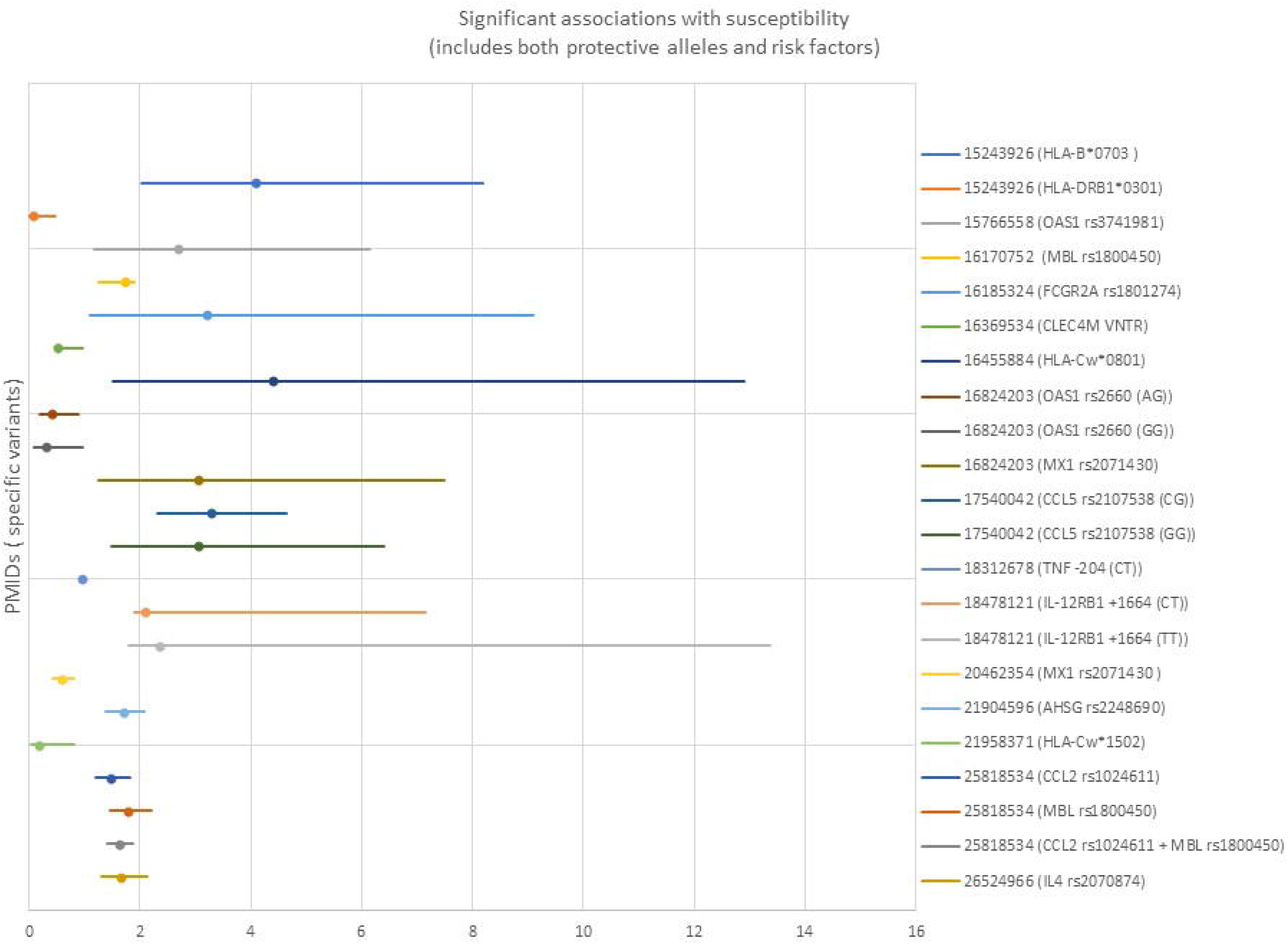
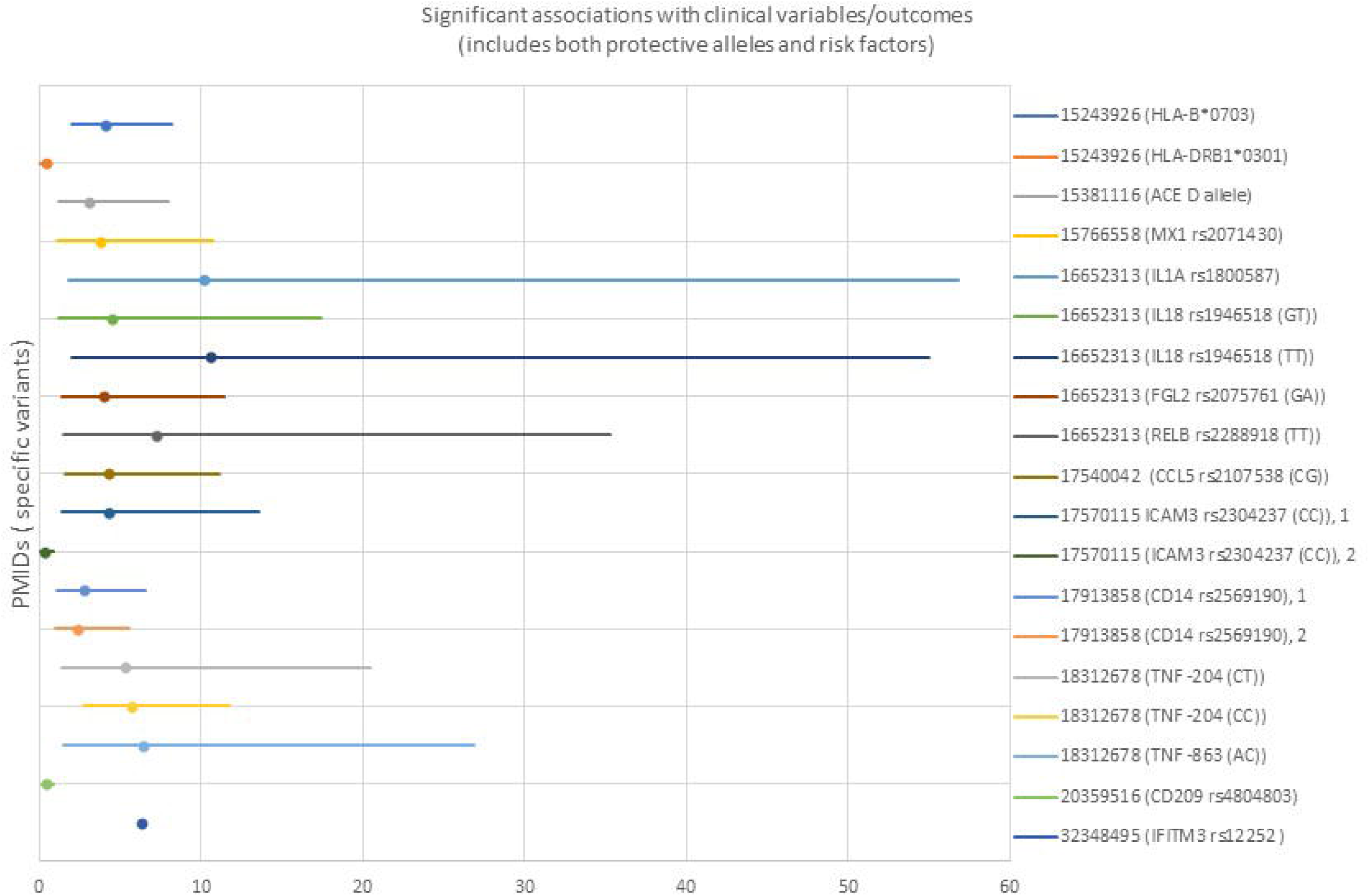
3A: Significant genetic associations with human susceptibility to coronavirus disease. Both protective and permissive genes are shown. Only studies reporting odds ratios (OR) and confidence intervals are shown. 3B: Significant genetic associations with human clinical variables and outcomes related to coronavirus disease. Both protective and permissive genes are shown. Only studies reporting odds ratios (OR) and confidence intervals (CI) are shown (PMID 32348495 did not include Cl).

Of the 35 human studies meeting the host genetic study criteria described above, 32 (91%) involved SARS-CoV-1, while 3 (9%) involved SARS-CoV-2. Two of the three SARS-CoV-2 studies were case reports (one on a single family, the other on two patients with a rare immunodeficiency) without specific studies related to host factors; it is anticipated that many more studies on SARS-CoV-2 will be published soon.

All of the association studies except one were candidate-gene analyses based on genes hypothesized to be important in disease susceptibility or clinical variables/outcome. The exception was a meta-analysis of 386 studies on susceptibility to tuberculosis, influenza, respiratory syncytial virus, SARS-CoV-1, and pneumonia.^174^

Candidate studies ranged from studies of single variants to studies of over 50 genes selected due to biological plausibility; seven of these studies focused on HLA alleles. Sixteen significant loci related to susceptibility to coronavirus were reported (of which 7 identified protective alleles). Sixteen significant loci related to outcomes or clinical variables were reported (of which 3 identified protective alleles). The types of cases and controls used varied. Only four studies used separate cohorts for replication/validation. However, the studies used many different types of cases and controls, including within the same study. For example, some studies compared healthcare workers with SARS-CoV-1 infection with healthcare workers who tested negative. Others compared data from individuals with documented infection with data from control samples taken from blood donors. Four studies conducted laboratory-based biological studies in addition to association analyses. These studies are summarized in Table 1 and Figure 3; more details are available in Supplementary Table 2.

### Additional papers on humans and other species

As described, human and animal studies have examined various host factors related to coronavirus infection. For example, human^175^ and animal^33,52,125^ studies have implicated age as having significant associations with outcomes; age appears to be strongly correlated with COVID-19 outcomes.^176^ The overall explanations remain unclear, but could at least partially involve age-related gene expression. Sex also appears to have a role. Human studies of SARS-CoV-1 and SARS-CoV2 suggest a correlation between sex and certain clinical parameters, perhaps rooted in sex-based or related immunologic differences.^175,177,178^ However, separating biological differences from sex-related cultural practices (e.g., different rates of social distancing) may be difficult.

Animal studies also suggest sex effects in multiple species, such as related to disease severity.^57,179^ Multiple studies examined different genes/proteins to determine disease susceptibility, transmissibility, and pathogenesis in various species. In addition to humanized genes, such as used in mouse models, studies have involved a combination of computational and biological approaches, and have investigated the viral entry receptors *ACE2* in SARS-CoV-1^10,35,157,158,180–183^ and SARS-CoV-2^184^ (for which there already exists a large body of unpublished and preprint work) and *DPP4* in MERS.^71,148,185–188^ Among other findings, these studies examined specific protein residues that are critical in viral-host interactions [18448527],^157^ Other studies examined manipulations of various genes/proteins to study the functional biological effects, including of *ANPEP,^189^ GLTSCR2,^190^ IFITM1, IFITM2*, and *IFITM3,^191^* and *MAVS.^192^*

## Discussion: human studies

Traditional genome-wide methods have been applied to human viral infections generally,^174,193^ but results have not been specific to coronaviruses, and it is unclear to what extent the observations are relevant to the current pandemic. Several dozen studies have investigated human genetic factors related to coronavirus infection. However, these studies have been limited by several potential factors. For endemic human coronaviruses, the mildness of disease may have deprioritized these studies; similar observations may explain the relative dearth of serologic knowledge related to these pathogens.^6^ For coronaviruses associated with more severe human disease, such as MERS and SARS-CoV-1, the fact that these epidemics were limited more than the current pandemic crisis may have fortunately led to a lack of cases with which one might conduct traditional association studies (unlike some other respiratory infections leading to more widespread disease).^194,195^ Additionally, these two severe conditions primarily affected human populations prior to the technological developments that led to wide availability of much cheaper and faster genomic sequencing.^196^

As shown (Supplementary Table 2), the small sample sizes of previous studies may have led to the preponderance of candidate gene studies. The sample sizes may also have precluded significant findings due to limitations of statistical power and the ability to replicate or validate findings. As previous human studies occurred in areas of the world affected by the coronavirus studied, it is possible that results from these studies would not extrapolate to other populations. Finally, different genes and loci are involved than those previously hypothesized. That is, hypothesis-free approaches may identify significant loci that were not identified by candidate approaches.

Based on announcements about multiple large-scale projects on host genetic factors and SARS-CoV-2, as well as the existence of larger genomic datasets that can be mined quickly and new methods that can be used to address biological questions,^197^ it is anticipated that considerable efforts - and an unfortunately large pool of research subjects - and may yield significant new results quickly.

### Limitations

There are multiple limitations to our summaries and analyses. First, it is likely that relevant articles were missed by our search process, and that key findings - including the study of certain genes - were therefore omitted. Along these lines, important findings within identified articles may also have been missed. Second, this analysis focused on DNA-based variants. These DNA-based genetic changes include those studied and identified through association studies as well as genes that were manipulated in experimental approaches, such as via knockout models to understand disease pathogenesis. Related ‘omic approaches, such as targeted or broad transcriptomic or proteomic studies, are frequently used to understand important aspects of disease. These approaches can lead to knowledge regarding specific genetic changes. For example, observed transcriptomic changes may enable the identification of important DNA-based variants that explain disease by correlating transcriptomic data with results of DNA sequencing.^198^ However, we categorized non-DNA based ‘omic approaches separately from DNA-based studies, and did not attempt to comprehensively recapitulate what is known about host reaction to disease. Finally, as the studies varied in many aspects, such as how cases and controls were defined, and which loci were interrogated, we were careful about comparing or combining data between different studies.

## Data Availability

All data referred to in the manuscript are available either in the main manuscript or in supporting files.

## Acknowledgments

This research was supported [in part] by the Intramural Research Program of the National Human Genome Research Institute, National Institutes of Health. PD was supported by the US National Institutes of Health award number 2R01AI148049–21A1. DATC was supported by the US National Institutes of Health award number R01-AI114703–01.

## Notes

### Competing Interest Statement

BDS was previously an employee of Opko Health, which includes subsidiaries that conduct testing relevant to COVID-19 and genetic/genomic testing.

### Clinical Trial

N/A

### Funding Statement

DATC was supported by the US National Institutes of Health award number R01-AI114703-01.
PD was supported by the US National Institutes of Health award number 2R01AI148049-21A1.

### Author Declarations

No IRB and/or ethics committee approvals were required for a systematic review of the literature.

